# A Platform for Data-centric, Continuous Epidemiological Analyses

**DOI:** 10.1101/2022.04.19.22274026

**Authors:** Flávio Codeço Coelho, Daniel Câmara, Eduardo Araújo, Lucas Bianchi, Ivan Ogasawara, Jyoti Dalal, Ananthu James, Jessica Lee Abbate, Aziza Merzouki, Izabel Reis, Kenechukwu Nwosu, Olivia Keiser

## Abstract

Guaranteeing durability, provenance, accessibility, and trust in open datasets can be challenging for researchers and organizations that rely on public repositories of data critical to epidemiology and other health analytics. Not only are the required repositories sometimes difficult to locate, and nearly always require conversion into a compatible format, they may move or change unpredictably. Any single change of the rules in one repository can hinder updating of a public dashboard reliant on pulling data from external sources. These concerns are particularly challenging at the international level, because systems aimed at harmonizing health and related data are typically dictated by national governments to serve their individual needs. In this paper, we introduce a comprehensive public health data platform, the EpiGraphHub, that aims to provide a single interoperable repository for open health and related data, curated by the international research community, which allows secure local integration of sensitive databases whilst facilitating the development of data-driven applications and reports for decision-makers. The platform development is co-funded by the World Health Organization and is fully open-source to maximize its value for large-scale public health studies.

## Introduction

Two years into the COVID-19 pandemic, the global health community has had to overcome new and unexpected challenges. Perhaps the biggest challenge beyond the immediate need for materials to protect against transmission and to test and treat patients has been getting timely and informative evidence into the hands of authorities for making effective public health decisions(1). The free flow of scientific and statistically accurate data, taking the form of clinical and laboratory information about the epidemic, remains a challenge even for resource rich countries. This ongoing challenge relates to the efficient and timely flow of such information, for the purpose of supporting an effective fight against viral spread (2–4). The difficulty in dealing with this technical data tsunami comes from many factors:

1. Slow infrastructure for reporting of health data. Prior to this pandemic, sharing of disease surveillance data was often slow, taking weeks or months (5,6) to reach country-level data repositories. COVID-19’s Omicron variant had spread to 89 countries within approximately one month of being detected (7).
2. Insufficient testing capacity. Since the beginning of the pandemic, production of test kits increased substantially, but the costs and the logistics for large-scale testing is still beyond reach for many countries(8).
3. Lack of a common global data model for disease reporting. Comparability of surveillance datasets is paramount to manage risks at a global scale (9,10).
4. Lack of interoperability for data exchange between countries.

Improving items 1 and 2 depends strictly on country funded infrastructure, but the last two items can be tackled through concerted actions by non-governmental actors and the global health research community in general(11). The development of an ecosystem of tools for online analysis of the surveillance data-stream creates a demand for better quality primary data, which can act as an incentive for countries to invest more in health data monitoring infrastructure.

In this paper, we present an initiative co-funded by the World Health Organization to build an open source platform for continuous epidemiological data analysis. It aims at filling multiple gaps in the current public-health data analysis ecosystem whilst functioning as a simple-to-use and responsive tool for decision-makers. We call this platform EpiGraphHub. It provides for automated data integration, cleaning, and harmonization, combined with a web interface for easy data exploration and building of live, interactive dashboards.

The proposed platform shares some similarities with other open data platforms as shown in Table 1, however it differentiates itself for the additional features it provides as well as for its focus on public health data and epidemiological data analysis.

**Table 1:** Feature comparison between EpiGraphHub and other open data platform tools. This comparison is not meant to be a complete evaluation of other tools, just a comparison on selected features of EpiGraphHub. Legend: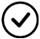 : yes,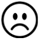 : No and 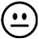: partially.

| Feature | *EpiGraphHub* | Our World in Data | CKAN(12) | Socrata | Google Data Studio |
| --- | --- | --- | --- | --- | --- |
| Automated, user-defined data collection | ☑ | ☹ | ☹ | ☹ | ☹ |
| Connects to external databases | ☑ | ☹ | 😐<sub>1</sub> | 😐<sub>1</sub> | ☑ |
| Dashboard creation | ☑ | ☹ | 😐<sub>2</sub> | 😐<sub>2</sub> | ☑ |
| Codeless use | ☑ | ☑ | ☑ | ☑ | ☑ |
| User-specific data processing and analysis | ☑ | ☹ | ☑ | ☑ | 😐 |
| Open Source | ☑ | 😐 | ☑ | ☑ | ☹ |
| Cloud or local deployment | ☑ | ☹ | ☑ | ☑ | 😐 |
| Integrated SQL IDE | ☑ | ☹ | ☹ | ☹ | ☑ |
| Epidemiological analysis libraries | ☑ | ☹ | ☹ | ☹ | ☹ |

1 https://epigraphhub.org
2 https://gadm.org/

The remainder of the paper is divided in 7 sections, *System Architecture* - where the architecture of the platform is described along with its design goals; *Data Collection* - in this section the general design of the data collection module is detailed; *Data Transformation* - where the set of transformations available to clean and preprocess datasets are described; *Data Exploration and Visualization* - The interactive web interface for visual analytics is presented in this section; *Data Analyses* - Here the Analytical methods currently implemented are described with examples; *Application Hosting* - In this section, we describe the usability of the EpiGraphHub platform as a common backend for web and mobile applications; Finally, the *Discussion* section talks about the platform’s relevance and applicability in the context of similar initiatives.

In order to make such continuous analyses possible, data from multiple sources must be integrated into a consistent data model before it can be used for analyses. A local storage layer is also provided to guarantee the persistence of the data used in the analyses. Moreover, for dynamic datasets (that are updated regularly), snapshots can be created, so the exact version of a dataset associated with published analyses can also be persisted.

## Methods

### System Architecture

EpiGraphHub is built upon the open source business intelligence platform Apache Superset(13), with added features to make it more suitable to epidemiological data analysis. As such, it is already able to scale well horizontally, to serve heavy loads on a distributed computing infrastructure.

The architecture of the platform facilitates the automated collection, transformation, storage, and visualization of data on an entirely open-source software stack. Portability and replication of the entire stack is an important aspect of the platform’s design, which places each software service within Docker containers (Figure 2).

**Figure 1:**
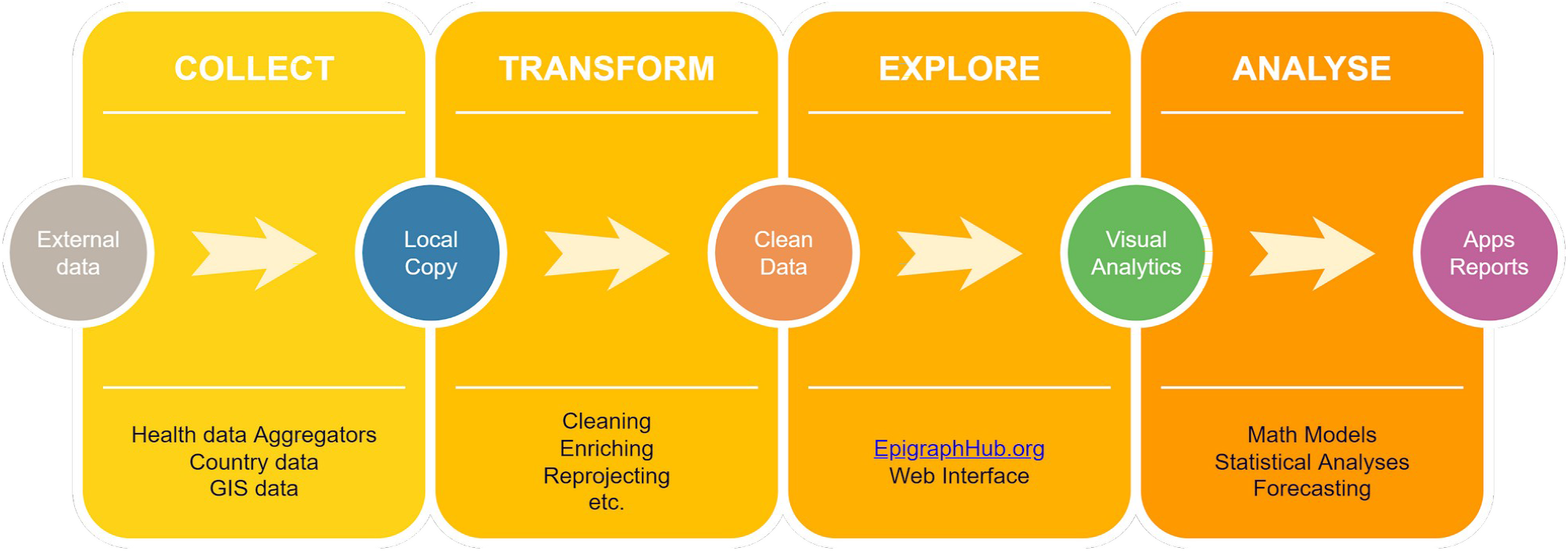
Overall organization of the EpiGraphHub platform, with its four main modules.

**Figure 2:**
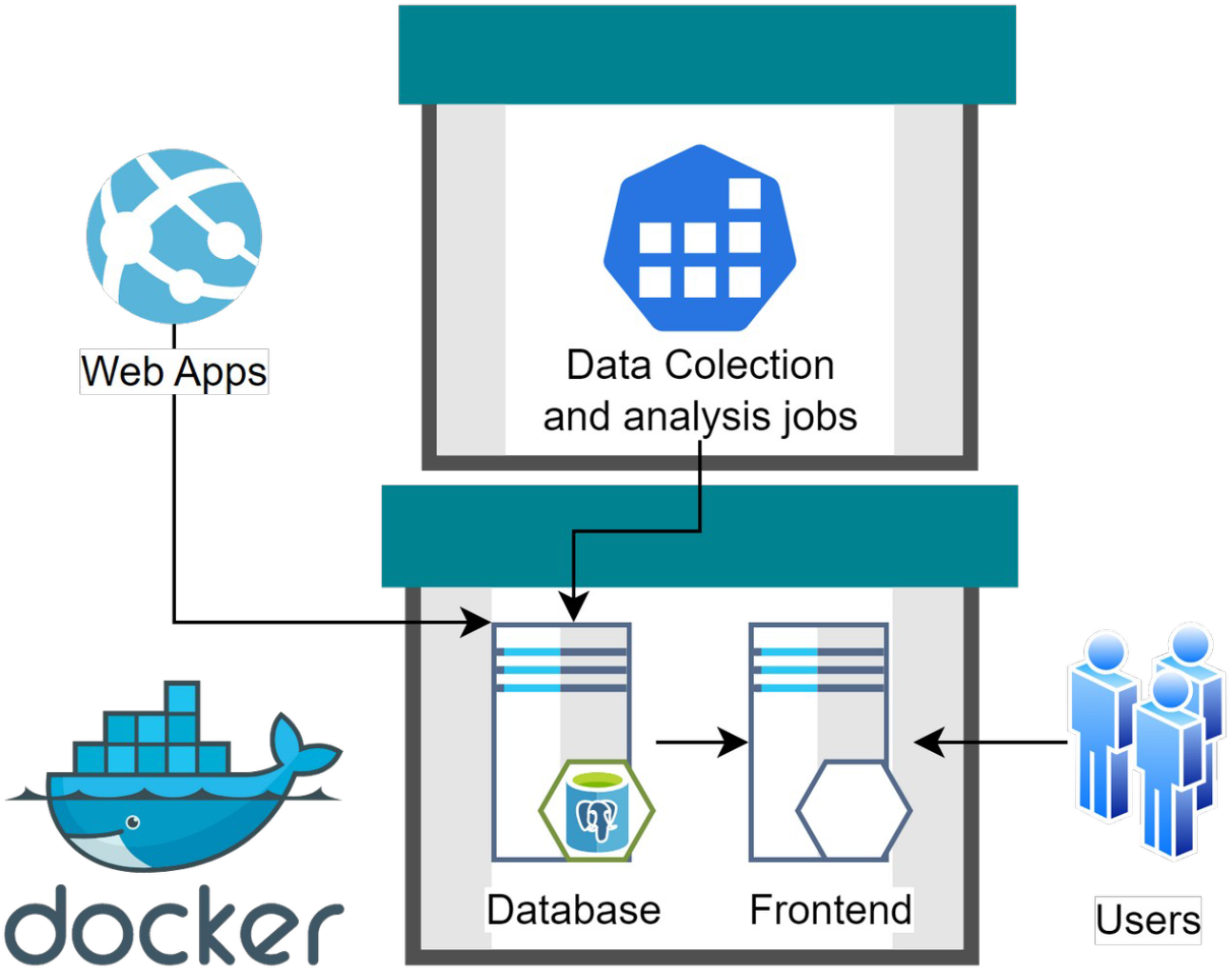
Platform architecture is based on docker containers. A container is a standard unit of software that packages up code and all its dependencies, so the application can be deployed quickly and runs reliably across different computing environments. In the diagram above, two of the key components of the platform are represented as the boxes with green lids. Containers are connected into a virtual network within the host environment and can exchange data. External web apps can connect directly to the containers to request data via the exposed APIs.

To maximize code re-usability and value for the open source community, all the tools for data collection, transformation and analysis are developed as EpiGraphHub software libraries available both in Python and the R languages. These libraries are fully documented, and ready for usage independent of the platform as well. Figure 3 show how a simple data upload to the platform can be accomplished with Python code.

**Figure 3:**
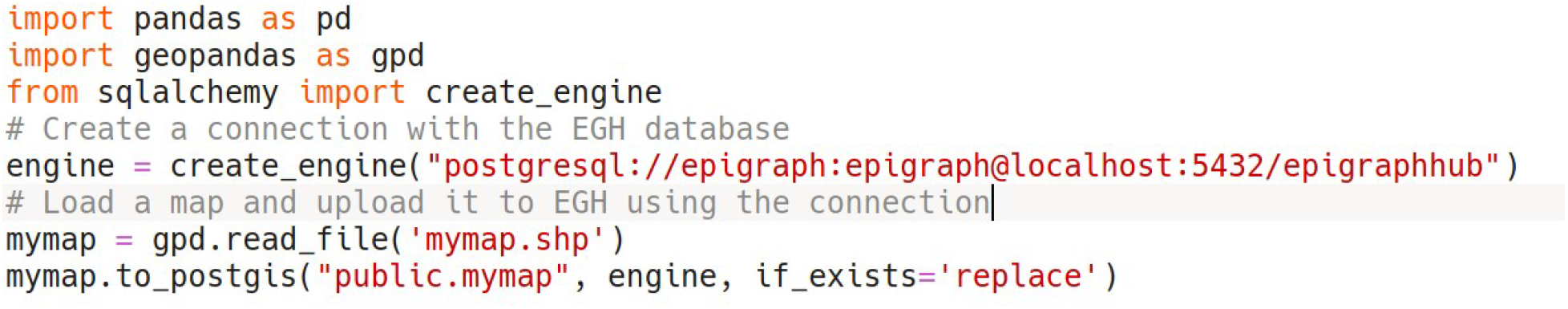
Python script to upload data to EGH, Once an encrypted SSH connection has been established.

The project provides a continuous integration (CI) workflow, based on the GitHub actions tools, CI consists in pre-configured testing scripts that run before any contribution can be merged into the main branch of our repositories. The CI routine builds all containers, checking for any misconfigurations and run unit tests on the EpigraphHub libraries. CI is key to preventing contributions from breaking the platform. All pull-requests have to pass CI tests before being merged into the main branch.

A continuous deployment (CD) workflow is currently under development to enable new releases on GitHub to automatically trigger an update on specified deployment servers. The CD workflow is an important tool to allow for efficient delivery of new features to users.

## Data Collection

There are many openly accessible repositories of health data (14–16). One of the key challenges of building a data analysis platform onto a broad set of data sources, which operate under distinct governance systems and are funded by a variety of different sources, is the long-term availability and immutability of these datasets. Lin et al.(17) defines the TRUST principles for open datasets: **T**ransparency, **R**esponsibility, **U**ser focus, **S**ustainability and **T**echnology. Another set of principles, known as the FAIR data principles (18) mention that all open data repositories should be **F**indable, **A**ccessible, **I**nteroperable, and **R**eusable. The platform follows all these principles, but also guarantees that all the data used for a particular analysis remains fixed and accessible as it was at the time the analysis was done.

In order to allow for that, we keep a copy of the original data in our servers, whenever its licensing terms permit. In the implementation of this replication, we make a distinction between static and dynamic datasets. Static datasets are the ones not subject to updating and revision – these datasets are imported only once. Dynamic datasets, on the other hand, must be periodically updated or extended. A good example of these are disease surveillance databases such as COVID-19 case counts. For these datasets, we define a periodicity for their updating, which is automatically triggered by the platform. All the workflow related to the data collection and integration is implemented using Apache airflow. Apache airflow is a distributed task scheduler, that allow for computational tasks defined as parts of a workflow DAG to be efficiently scheduled and monitored.

All datasets, raw or transformed, are stored into a PostgreSQL relational database. On this database server, data is organized according to access level, wherein public and restricted access datasets are kept on entirely separate databases (figure 4). This guarantees that we can expose the full contents of the public datasets on our web-based visual analytics tool, without compromising the security of the non-public datasets.

**Figure 4:**
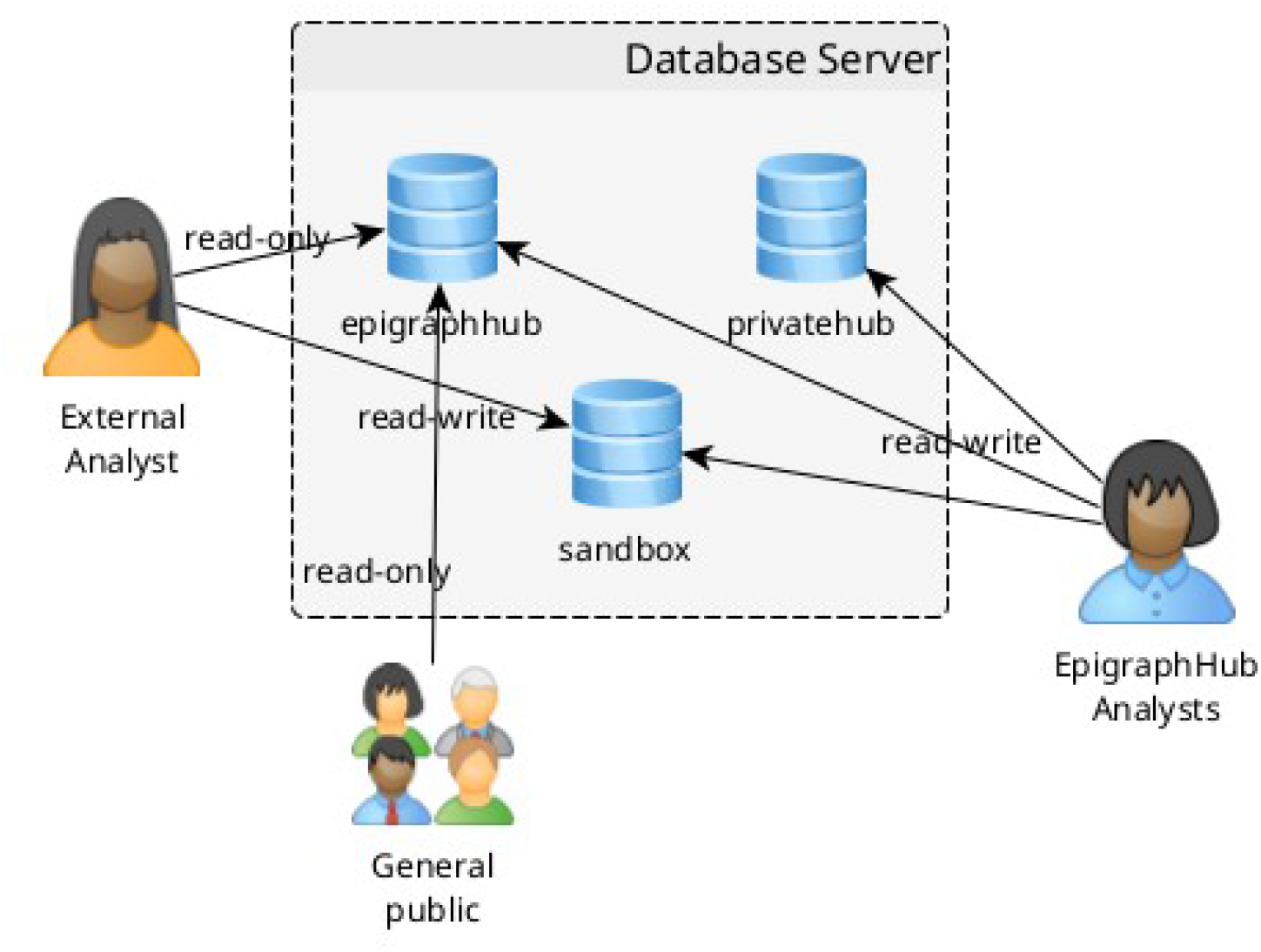
Organization of the datasets within the EpiGraphHub database server. Access control to the datasets can be configured according to the requirements of each dataset.

One key aspect of the platform is its ability to connect and pull data from widely used health information management systems (HIMS) such as DHIS2, GoData and others. Governmental and also private HIMS are typically closed platforms, where only authorized personnel have access to the data. Therefore, EpiGraphHub aims to make available open source software to make it as easy as possible to pull data from these platforms, given the required authorization from data owners. Our cloud service at EpigraphHub.org, also includes an integrated deployment of the also open source Kobo-toolbox server, provided free of charge to partners that need to collect primary data.

### Data Transformation

As an integral part of the Data integration pipeline, some datasets may require some transformations applied before they are inserted into the database for storage. Examples of such transformation can go from simple normalization of dates to a standard, Addition of ISO geocodes to facilitate the construction of maps, etc. These transformations are designed to not alter in any way the semantics of the data, or its scope. They focus rather on enriching it, or making it consistent to data representation standards. As data collection and transformation scripts document the source of the original data and the transformations applied, all transformations are clear to users, who are able to modify them for their personal use.

### Data Exploration and Visualization

A web interface for interactive querying and visualization of the platform’s datasets is available^3^, to help users without programming skills to create visualizations from data queries. The results from the queries can then be published or shared or be integrated into live dashboards that get updated whenever the underlying data changes. This interface is based on the open source visual analytics platform Apache Superset. Figure 5 shows a dashboard created on the platform.

**Figure 5:**
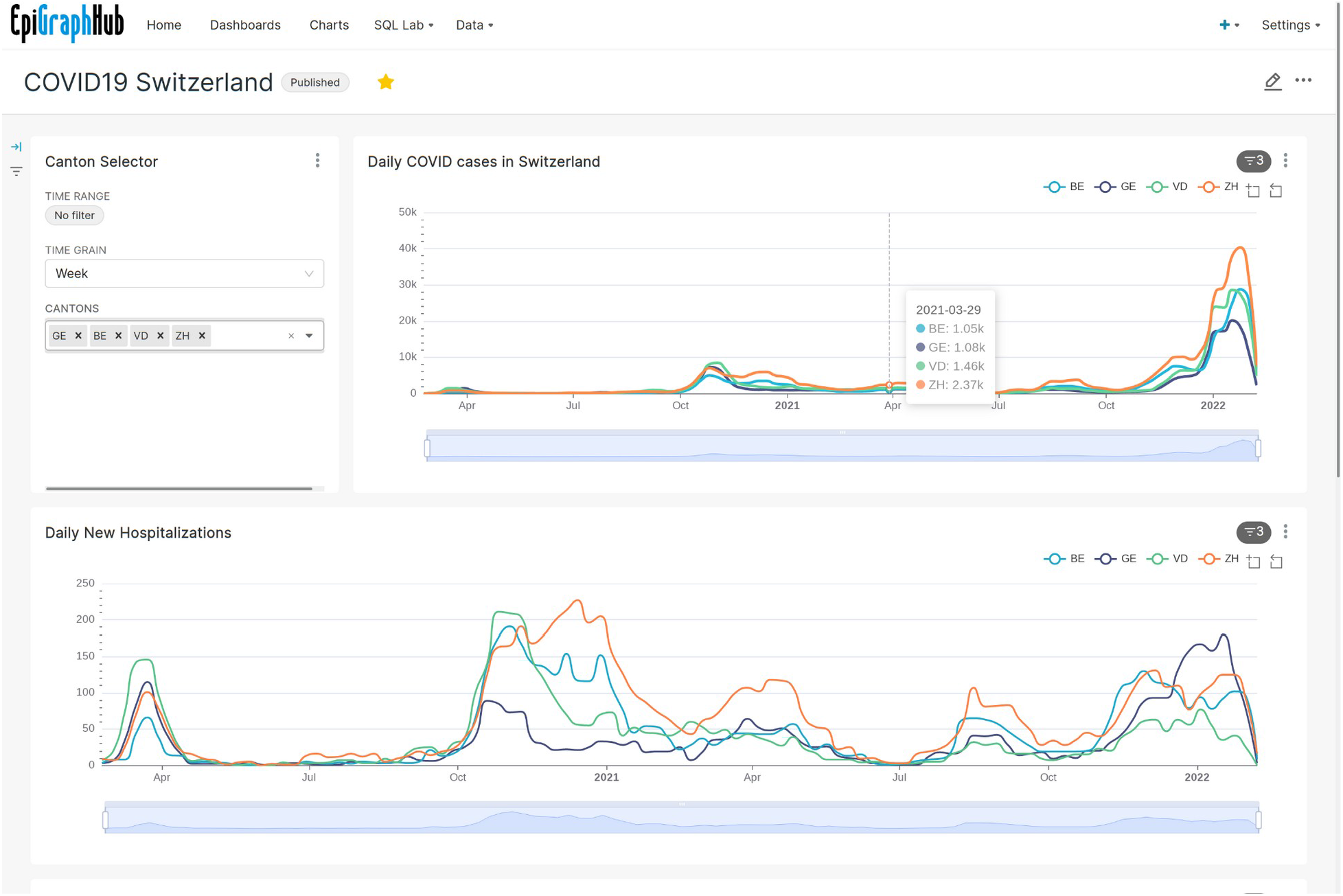
Dashboard created entirely by pointing and clicking on the EpiGraphHub platform, Showing COVID-19 cases and hospitalizations in Switzerland by canton.

Sharing and publishing of the visualizations built inside the platform can be done in multiple ways. All charts, SQL Queries, and Dashboards created and saved by a user are given a permanent URL which can be shared with users outside the platform. Dashboards can be flagged as published, meaning they can be seen by all users.

This web platform allows for fine-grained access control to datasets via user permissions that can be customized for users that create an account on the platform.

Another advantage of the EpiGraphHub Web interface is that it offers an accessible environment for a gentle introduction to SQL (Structured Query Language) through the possibility of translating point-and-click based queries performed on the platform into the equivalent SQL code which can be saved, shared and further modified by users.

### Data Analyses

Different levels of analyses are possible within the EpiGraphHub platform. One level is the data cleaning and transformation procedures which are applied between the data collection and its storage into the platform’s database. Then we have the analyses which are done within the database through the web interface either through the graphical interface or by writing arbitrary SQL code that runs on the server. Finally, we have the more advanced, analytical API, exposed through the EpiGraphHub Python and R Libraries. These Software libraries expose a number of analytical methods as well as data access functions that help users to create their own analytical applications based on EpiGraphHub’s available data.

One example of a data analysis workflow within the EpiGraphHub platform was the support to the African regional office of the World Health Organization (WHO AFRO) during the first year of the COVID-19 pandemic (19,20). Individual case data from member states was received as national linelists stored as Excel® files. Data quality checks were performed for each country linelist, and datasets were then harmonized to have standardized variables. The following step was the integration of all datasets into the EpigraphHub database using our libraries. These datasets contained individual level information for all countries. All variables related to the geographical localization of the cases (such as residence, local of infection, local of reporting, among others) were then standardized using ISO 3166-1 alpha-3 codes for country, district and province levels. These harmonization procedures were then followed by integration with data provided by the GADM project^4^ for all three administrative levels. Population data was obtained via raster files from the Gridded Population of the World (GPW v4^5^) collection, which provided fine resolution for population counts inside districts and provinces of the analysed countries. All these steps of data harmonization were then followed up by the development of weekly and monthly analytical reports, which included exploratory epidemiological analyses. The datasets for this project cannot be shared publicly, But that does not reduce the utility of the EpiGraphHub platform as it has been designed to host both public and restricted dataset as already described above. This allows analysts with the proper permissions to combine both public and private datasets on the same analytical report or dashboard.

### Application Hosting

The EpiGraphHub also provides a workflow for quick deployment of Web or mobile applications based on the platform. This workflow allows for continuous integration and deployment from a GitHub repository. It currently supports Python-based Streamlit® and R-based Shiny® applications. Other application frameworks may be supported in the future.

Applications are encapsulated within docker containers hosted on our server. This allows for low latency in accessing Hub-hosted datasets. Docker templates are available for app developers to test and deploy their apps locally, without having to bother with deployment details. Applications in production are updated via a continuous deployment workflow that allows for a rebuild of the containers whenever a new release is created in its GitHub repository. This methodology facilitates the deployment of multiple applications without overloading the platform administrators.

### Patient and Public Involvement

This work was carried on without the involvement of patients or the general public.

## Discussion

A number of health data aggregation platforms have been developed in recent years, particularly in response to the COVID-19 pandemic. The problem is not new. In Brazil, the Infodengue and Infogripe projects have been monitoring arboviruses and Influenza respectively and making data and epidemiological analyses open to the general public for many years (21,22). Hürliman et al. (23), in 2011, talks about the need for a global database to monitor neglected tropical diseases, but more than ten years later no such database exists.

During the COVID-19 pandemic a number of new data repositories for disease surveillance have been developed. For example, *Our World in Data (OWID)*, spearheaded by Oxford University, not only focused on disease surveillance, has played an essential role during the pandemic to guarantee an openly accessible global perspective of the global spread of SARS- CoV2 and its variants (24). OWID also maintains live dashboards for interactive exploration of their datasets. Google also entered this arena, through its public datasets program with the construction of a fine-grained collection of COVID19 data (15). Google’s datasets excel on ease of use, and built-in functionality for quick visual explorations of the data.

The COVID19 pandemic has also spurred many countries to make their own COVID19 data available on the web. However, we suspect that this attitude will not survive the end of the pandemic. The best indication of this is the fact that the openness observed for COVID data has not yet been extended to other communicable diseases. The need for more widespread transparency towards data that is of interest to public health is clear. Such transparency cannot come only eventually, in response to visible health emergencies. Rather, it must be an integral part of governmental infrastructure for health data management.

The data Integration aspect of EpiGraphHub goes further than helping with openness, but it also brings together all the essential pieces of the data puzzle of effective public health policies(25). Moreover, being a completely open source package that can easily be deployed in a country, it fills a gap with an easy to deploy and low-cost solution for health data analysis.

The EpiGraphHub platform, although still in development, is already being used to help in the response to the COVID-19 pandemic in many countries(19,20). As the pressure for urgent responses to the current pandemic finally wanes, we hope our platform will be ready to continue to provide value to disease surveillance programs across the globe and serve as a prime example of the benefit of openly accessible public health data.

## Data Availability

All data produced are available online at

https://epigraphhub.org

## Footnotes

3 https://sedac.ciesin.columbia.edu/data/collection/gpw-v4

4 Can create “data preview” charts

5 Can federate across instances

